# Greener Operations: a James Lind Alliance Priority Setting Partnership on environmentally sustainable peri-operative practice

**DOI:** 10.1101/2022.07.12.22277545

**Authors:** Max Clayton-Smith, Hrishi Narayanan, Clifford Shelton, Louise Bates, Fiona Brennan, Beck Deido, Mike Donnellon, Jennifer Dorey, Bob Evans, Jonathan Gower, Yasmina Hamdaoui, John Hitchman, Stephen Michael Kinsella, Rebecca Knagg, Cathy Lawson, Daniel Morris, Victoria Pegna, Tracey Radcliffe, Olivia Schaff, Tim Sheppard, Jennifer Strong, David Jones

## Abstract

**Objectives:** To agree the ‘top 10’ research priorities for environmentally sustainable peri-operative practice.

**Design:** surveys and literature review; final consensus workshop using a nominal group technique. Setting: UK-based.

**Participants:** healthcare professionals, patients, carers, and the public.

**Outcome measures:** initial survey- suggested research questions; interim survey- shortlist of ‘indicative’ questions (the 20 most frequently nominated by patients, carers and the public, and healthcare professionals); final workshop- ranked research priorities.

**Results:** initial survey- 1,926 suggestions by 296 respondents, refined into 60 indicative questions. Interim survey- 325 respondents. Final workshop- 21 participants agreed the ‘top 10’:

- How can more sustainable reusable equipment safely be used during and around the time of an operation?
- How can healthcare organisations more sustainably procure (obtain) medicines, equipment and items used during and around the time of an operation?
- How can healthcare professionals who deliver care during and around the time of an operation be encouraged to adopt sustainable actions in practice?
- Can more efficient use of operating theatres and associated practices reduce the environmental impact of operations?
- How can the amount of waste generated during and around the time of an operation be minimised?
- How do we measure and compare the short- and long-term environmental impacts of surgical and non-surgical treatments for the same condition?
- What is the environmental impact of different anaesthetic techniques (e.g., different types of general, regional and local anaesthesia) used for the same operation?
- How should the environmental impact of an operation be weighed against its clinical outcomes and financial costs?
- How can environmental sustainability be incorporated into the organisational management of operating theatres?
- What are the most sustainable forms of effective infection prevention and control used around the time of an operation (e.g., PPE, drapes, clean air ventilation)?

**Conclusions:** a broad range of ‘end-users’ have identified research priorities for sustainable peri-operative care.

**Strengths and Limitations of this Study:**

- We have defined the top 10 research priorities according to healthcare professionals, patients, carers, and members of the public, in an important and expanding area of health research.
- To our knowledge, this is the first research priority setting partnership in any field of sustainable healthcare.
- Patients, carers, and members of the public comprised 21% of survey respondents overall, a smaller proportion than in many priority setting partnerships. This may have been because of the online methods used (due in part to the COVID-19 pandemic) or the novel subject matter.
- The James Lind Alliance process is consensus-based, transparent, and includes measures to ensure that patient, carer and public opinions are represented.
- The scope of our work was limited to ‘care provided from or in the secondary care setting to patients who may benefit from surgical management’ so does not include the full patient journey; more sustainability-focussed priority setting partnerships would be beneficial in the future.

**Original Protocol of the Study:** See https://www.jla.nihr.ac.uk/documents/greener-operations-sustainable-peri-operative-practice-psp-protocol/27106

## Introduction

It is increasingly recognised that healthcare, as a resource-intensive industry, makes a significant contribution to environmental harms such as global warming and pollution.^1^ In turn, these environmental harms contribute to ill health, thereby creating an increased demand for healthcare services.^2^ In the UK, National Health Service trusts are recognised as an ‘anchor institutions’, large organisations that influence the health and wellbeing of their communities not only through providing healthcare – but though practices in procurement, employment, community engagement and environmental responsibility.^3^ Recently, healthcare systems, educational regulators and professional societies have begun to issue guidelines and implement plans aiming to mitigate the carbon footprint and ecological impacts of healthcare.^4-9^ This rapid expansion of interest in the area is both necessary and welcome but presents its own challenges. Though there are several high-impact measures that should be urgently implemented (e.g., decarbonisation of electricity production),^4^ it is universally acknowledged that achieving sustainable healthcare will require research and innovation.^4-6^

Between 220 and 344 million operations are thought to be performed worldwide every year,^10^ a number which will increase as the Lancet Commission on Global Surgery target of 5,000 operations per 100,000 population is approached.^11^ The peri-operative journey, from initial consultation to surgery and then discharge from hospital and recovery, is a complex process that involves many groups of hospital staff. Operations are known to be among the most resource-intensive healthcare interventions;^12^ each operating theatre creates over two tonnes of solid waste per year,^13^ and a single operation can generate a ‘carbon footprint’ equivalent to driving more than 2,000 miles.^14^ Peri-operative practice therefore represents a significant opportunity to make healthcare more environmentally sustainable. This opportunity has not gone unrecognised, and recent years have seen a proliferation in research funding, fellowship posts, and publications relating to sustainability in the peri-operative period.^15-17^

Noting the increasing interest in research relating to sustainability in peri-operative practice, we felt that this represented an ideal subject for a James Lind Alliance (JLA) Priority Setting Partnership (PSP), in order to direct and inform future research.

The JLA is a not-for-profit organisation, founded to address evidence uncertainties in specific areas of research through collaboration between patients, carers and clinicians.^18^ Using an ‘open-to all’ survey-based approach the JLA seeks to engage the ‘end users’ of research to help direct funding to the areas of greatest need, thereby minimising biases caused by financial or purely scientific research motives. Since its founding in 2004, it has facilitated more than 140 PSPs, developing a robust methodology to identify the ‘top 10’ research priorities in a given subject area.^19^

In 2019, we were successful in our application to the JLA to run ‘Greener Operations’, a PSP which aimed to identify the top 10 unanswered research questions connected to environmentally sustainable peri-operative practice, as defined by an expansive group of patients, carers, members of the public and healthcare workers. We believe this to be the first PSP to be conducted in any field of sustainable healthcare.

## Methods

The Greener Operations PSP was conducted according to the standard JLA methodology by a team comprising project leads, information specialists, a multidisciplinary steering group composed of healthcare professionals and patient and public representative, and a James Lind Alliance advisor.^20^ The PSP was supported by partner organisations involved or interested in peri-operative care, such as professional associations, royal colleges, and patient groups. As this was a survey-based project publicly available to all on a voluntary basis, research ethics committee approval was not required.^20^ Potential participants were provided with an explanation of what each phase of the project involved, including how the data would be used, as described below.

### Setting up the priority setting partnership

Following approval of charitable funding, the PSP was established in August 2020 by the project leads. Two information specialists were appointed, to be responsible for managing the surveys and data analysis, and an advisor was assigned by the JLA. Partner organisations, responsible for promoting the PSP and ensuring that surveys reached as wide an audience as possible, were recruited by the project leads by email contact with organisational representatives. The steering group was formed by inviting expressions of interest from individuals linked to the partner organisations (e.g., members of environmental or peri-operative committees or working groups). We aimed to recruit a wide range of healthcare professionals involved in peri-operative practice, including surgeons, anaesthetists, nurses, operating department practitioners and pharmacists. In addition, the steering group included non-clinical professionals involved in sustainability, and individuals with lived experience of undergoing surgery who could represent patients’ interests.

Because of ongoing COVID-19 pandemic and a desire to minimise the environmental impacts of the project itself, it was agreed by the steering group that all meetings would be held online. The meetings were chaired by the JLA advisor, and conducted using a video-conferencing platform (Zoom, Zoom Video Communications, Inc, San Jose, California, USA).

### Defining scope

At the initial meeting of the steering group, the study protocol and scope of the PSP were confirmed.^21^ Though we recognised that the complete perioperative journey often commences and ends in the community, for pragmatic reasons we defined ‘peri-operative practice’ as that provided from or in the secondary care setting to patients who may benefit from surgical management, including:

- pre-operative assessment and optimisation (e.g. pre-operative clinic)
- counselling and shared decision-making (including on decisions regarding the appropriateness of surgery, and different approaches to peri-operative management)
- pre and postoperative hospital care (including outpatient, ambulatory, virtual and inpatient care)
- intra-operative management (including surgical and anaesthetic techniques)
- both clinical (e.g. surgical and anaesthetic techniques) and non-clinical (e.g. energy, water, waste management and recycling) aspects
- the implementation of these practices within organisations and departments
- achieving positive peri-operative outcomes

Care beyond the early post-operative period (e.g. prolonged rehabilitation), broader aspects of environmental sustainability, and non-UK practice were excluded, along with questions relating to denying management of illness purely on the basis of environmental sustainability.

### Gathering uncertainties

An initial online survey (SurveyMonkey, Momentive, San Mateo, California, USA) was used to invite patients, carers, healthcare professionals, and members of the public to suggest evidence uncertainties connected with sustainable peri-operative care. Respondents were asked to state, via free-text boxes, what questions they felt needed to be answered by future research to help make peri-operative practice more environmentally sustainable. To help respondents to consider the full scope of the peri-operative patient journey, we asked them to consider the pre-, intra- and postoperative phases, and also invited any further suggestions. In addition to suggested questions, demographic data were collected. After a pilot within the steering group, the initial survey was launched online on 10^th^ May 2021, and disseminated through partner organisations, the project website, and social media, using a web link and quick response (QR) code. Demographic data were routinely reviewed to consider whether the survey was successfully reaching all stakeholder groups. The survey remained open for 17 weeks, until 31^st^ August 2021.

### Data processing

After closing the survey, the raw data were downloaded for processing and analysis. To maintain data integrity and facilitate cross checking, each respondent was assigned a unique code number, with each individual response assigned a sub-code. Suggestions were assessed independently by the information specialists to determine whether they were in-scope or out-of-scope, based on the criteria in the PSP protocol. Where both information specialists agreed that a suggestion was out of scope, that suggestion was not analysed further. Suggestions that did not clearly fall in or out of scope were kept for further analysis, to ensure potentially relevant suggestions were not missed.

To aid with analysis, suggestions were categorised into themes by the information specialists based on subject matter. The themes and suggestions were then reviewed by members of the steering group to form a list of indicative questions, agreed by consensus. Suggestions that were deemed to be similar were combined to form a single indicative question; others that were deemed to be too broad were split into separate questions. Each in-scope suggestion was allocated to a minimum of one appropriate indicative question to ensure all data were kept in the analysis.^20^ The steering group then cross-checked the list of indicative questions with the individual suggestions to ensure that the meaning of the suggestions was captured appropriately.

### Literature review

A literature review was undertaken to identify if any of the indicative questions had already been answered by currently available research. Following standard JLA principles, questions were categorised as having been answered ‘completely’, ‘partially’, or ‘not at all’.^20^ For a question to be deemed ‘answered completely’, a relevant, up-to-date and reliable systematic review or national clinical guideline that addressed the question would be required. For each indicative question, we worked with our healthcare library to search relevant databases (EMBASE, CINAHL, Medline and the Cochrane Database of Systematic Reviews; see Appendix 1), and reviewed guidelines from the National Institute for Health and Care Excellence, the Scottish Intercollegiate Guidelines Network, and relevant Royal Colleges and professional associations. In addition, members of the steering group who were members of professional organisations checked if there were any guidelines ‘in press’. The findings of the literature review were reviewed by the steering group who decided whether any questions could be deemed to be answered completely. Questions that had some evidence available that did not meet the criteria for being completely answered were classified as partially answered, and these questions, along with the unanswered ones, were taken forward into the interim survey.

### Interim priority setting

A second online survey (SurveyMonkey, Momentive, San Mateo, California, USA) was used to rank the long list of indicative questions to generate a shortlist of the most important questions that could then be discussed at the final prioritisation workshop. Respondents were presented with the indicative questions, displayed in a random order unique to each respondent, and asked to select the 10 questions they felt were most important. In addition, routine demographic data were collected. The survey was open for six weeks from 19^th^ April to 30^th^ May 2022. Following this, the raw data were analysed to identify the questions selected the most frequently. To moderate the influence of unequal numbers of respondents from different backgrounds, equal weighting was given to questions selected by healthcare professionals, and those selected by respondents who classified themselves as patients, carers or members of the public. The 20 questions most frequently selected by these two groups were taken forward to the final priority setting workshop

### Final priority setting workshop

The final priority setting workshop was a one-day in-person event, conducted according to a nominal group technique,^22^ chaired and facilitated by a team of advisors from the JLA. Respondents to the interim survey were invited to express an interest in participating in the workshop, and invitations were made using a purposive approach in order to promote a balanced group in terms of background (healthcare professional or patient, carers or member of the public). Participants were asked, in advance, to consider the importance of the questions for discussion. The JLA advisors allocated participants to three small groups of up to 10 people, aiming for a diverse mix of backgrounds in each group. The process comprised five phases:^20^

1. Small group discussions: participants listed the three questions they felt were most important for research, and the three that they felt were least important. These were recorded by the facilitator, and an opportunity for further discussion and clarification was provided.
2. First round of small group ranking: in the same groups, the facilitator laid out the questions, printed on cards, in rough groupings: those which were thought to be most important by group members, those thought to be least important, and those not mentioned or where there was divergence of views. Group participants then prioritised all of the questions by moving the cards into rank order. The ranking scores of the interim survey were made available to participants at this phase, to assist with ranking decisions.
3. Plenary review: the ranking agreed by each group was entered into a spreadsheet, and assigned a value (highest rank = 1, second highest = 2, etc). These ranks were combined by addition to create an aggregate ranked list. The aggregate ranks were presented to all workshop participants in plenary, with an opportunity for discussion.
4. Second round of small group ranking: participants were allocated to new groups by the JLA advisors, aiming to maintain a balance of backgrounds and expose participants to a different range of views. These new groups discussed and revised the aggregate ranked list, again by moving cards positioned to reflect the rank order.
5. Final plenary review: as per phase 3, the small group scores were entered into a spreadsheet, and combined by addition. The aggregate ranking was presented to all workshop participants in plenary, with the cards laid out in order. The ranking was discussed in plenary group, in order to agree the final ranking.

The final workshop discussions were chaired by trained JLA advisers to ensure that no one group or individual dominated the decision making. The aim was to reach agreement by consensus at the end of each phase, with decisions made by majority vote if consensus could not be reached.

## Results

Two hundred and ninety-six individuals responded to the initial survey, of whom 230 (77.7%) classified themselves as healthcare professionals, 40 (13.5%) as members of the public, 21 (7.1%) as patients, and three (1.0%) as carers. Two (0.7%) did not state their background. Of the healthcare professionals, most were doctors (142; 61.7%), with nurses (23; 10%) and pharmacists (10; 4.3%) the next largest groups. Detailed demographics are displayed in table 1.

**Table 1:** demographic details of respondents to the initial and interim Greener Operations surveys

|  | Initial Survey | Interim Survey |
| --- | --- | --- |
| Total responses | 296 | 325 |
| **Gender** |  |  |
| Woman (including trans woman) | 171 (57.8%) | 187 (57.5%) |
| Man (including trans man) | 110 (37.2%) | 122 (37.5%) |
| Non-binary | 4 (1.4%) | 0 (0%) |
| Prefer not to say | 8 (2.7%) | 7 (2.2%) |
| Other | 1 (0.3%) | 4 (1.2%) |
| Question skipped | 2 (0.7%) | 5 (1.5%) |
| **Age** |  |  |
| Under 18 | 0 (0%) | 0 (0%) |
| 18-25 | 5 (17%) | 7 (2.2%) |
| 26-40 | 84 (28.6%) | 139 (42.8%) |
| 41-60 | 139 (47.3%) | 130 (40.0%) |
| 61-80 | 57 (19.4%) | 38 (11.7%) |
| Over 80 | 3 (1.0%) | 0 (0%) |
| Prefer not to say | 6 (2.0%) | 6 (1.9%) |
| Question skipped | 2 (0.7%) | 5 (1.5%) |
| **Ethnic group** |  |  |
| White | 230 (77.7%) | 258 (79.4%) |
| Asian or Asian British | 34 (11.5%) | 33 (10.1%) |
| Black, African Caribbean or Black British | 3 (1.0%) | 2 (0.6%) |
| Mixed or Multiple Ethnic Groups | 6 (2.0%) | 13 (4.0%) |
| Other ethnic group | 3 (1.0%) | 3 (0.9%) |
| Prefer not to say | 18 (6.1%) | 11 (3.4%) |
| Question skipped | 2 (0.7%) | 5 (1.5%) |
| **Region** |  |  |
| North West England | 82 (27.7%) | 115 (35.4%) |
| North East England | 35 (11.8%) | 31 (9.5%) |
| West Midlands | 12 (4.1%) | 21 (6.5%) |
| East Midlands | 17 (5.7%) | 14 (4.3%) |
| London | 33 (11.1%) | 28 (8.6%) |
| South West England | 18 (6.1%) | 19 (5.8%) |
| South East England | 52 (17.6%) | 43 (13.2%) |
| Scotland | 9 (3.0%) | 14 (4.3%) |
| Wales | 18 (6.1%) | 6 (1.8%) |
| Northern Ireland | 2 (0.7%) | 4 (1.2%) |
| Outside UK | 7 (2.4%) | 18 (5.5%) |
| Prefer not to say | 9 (3.0%) | 7 (2.2%) |
| Question skipped | 2 (0.7%) | 5 (1.5%) |
| **Background** |  |  |
| Patient | 21 (7.1%) | 19 (5.8%) |
| Carer | 3 (1.0%) | 2 (0.6%) |
| Member of the public | 40 (13.5%) | 45 (13.8%) |
| Healthcare Professional | 230 (77.7%) | 254 (78.2%) |
| Question skipped | 2 (0.7%) | 5 (1.5%) |
| **Profession (if healthcare professional)** |  |  |
| Advanced practitioner | 2 (0.9%) | 4 (1.6%) |
| Anaesthesia associate | 4 (1.7%) | 2 (0.8%) |
| Dentist | 2 (0.9%) | 2 (0.8%) |
| Doctor | 142 (61.7%) | 172 (67.7%) |
| Healthcare Assistant | 4 (1.7%) | 0 (0%) |
| Nurse | 23 (10.0%) | 16 (6.3%) |
| Operating department practitioner | 7 (3.0%) | 20 (7.9%) |
| Non-clinical role | 3 (1.3%) | 6 (2.4%) |
| Midwife | 0 (0%) | 4 (1.6%) |
| Paramedic | 1 (0.4%) | 0 (0%) |
| Perfusionist | 1 (0.4%) | 0 (0%) |
| Pharmacist | 10 (4.3%) | 1 (0.4%) |
| Physiotherapist | 2 (0.9%) | 0 (0%) |
| Porter | 1 (0.4%) | 0 (0%) |
| Radiographer | 0 (0%) | 1 (0.4%) |
| Other | 15 | 24 (9.4%) |
| Question skipped | 13 (5.7%) | 2 (0.8%) |

Respondents to the initial survey suggested 1,926 uncertainties for research. After initial review, we removed 309 suggestions agreed to be out-of-scope. After thematic categorisation of the remaining 1,617 suggestions, 78 themes were identified. This was further consolidated to 60 indicative questions by steering group consensus.

The literature review revealed that none of the indicative questions had been completely answered by currently available research. Members of the steering group who were part of professional bodies confirmed there were no relevant upcoming guidelines from their respective organisations that would answer the questions. Twenty-three questions were found to be partially answered by the available evidence. Therefore, all 60 indicative questions were included in the interim survey.

Three hundred and twenty-five individuals responded to the interim survey, of whom 254 (78.2%) classified themselves as healthcare professionals, 45 (13.8%) as members of the public, 19 (5.8%) as patients, and two (0.6%) as carers. Five (1.5%) did not state their background. Of the healthcare professionals, most were doctors (172; 67.7%), with operating department practitioners (20; 7.9%) and nurses (16; 6.3%) the next largest groups Detailed demographics are displayed in table 1.

The number of selections for each question was ranked separately according to whether respondents were healthcare professionals, or patients, carers and members of the public. A fractional ranking technique (tied ranks being assigned the mean of the ranking positions) was used to identify the 20 highest-ranked questions for each group, 14 of which were common to both groups (table 2). This led to 25 questions progressing into the final prioritisation workshop.

**Table 2:** highest-ranked indicative questions in the interim survey. Left column, healthcare professionals; mid column, patients, carers and members of the public; right column, combined rank. The top 20 questions for each group are highlighted in green. The combined rank (used for data organisation only) was calculated by adding the two rank scores, then ranking the added scores.

| Question | Healthcare Professional Interim Rank | Patient, Carer and Public Interim Rank | Combined Interim Rank |
| --- | --- | --- | --- |
| What alternative, more sustainable, materials can replace plastic packaging and equipment used during and around the time of an operation? | 1 | 1.5 | 1 |
| Can equipment be recycled or repaired, instead of being disposed of, to reduce its environmental impact? | 3 | 1.5 | 2 |
| How can the amount of waste generated during and around the time of an operation be minimised? | 2 | 3.5 | 3 |
| What are the most sustainable forms of effective infection prevention and control used around the time of an operation (e.g., PPE, drapes, clean air ventilation)? | 4 | 3.5 | 4 |
| How much recyclable waste generated during an operation is being appropriately recycled? | 5 | 5.5 | 5 |
| How do we define and avoid low-benefit or unnecessary operations? | 12 | 7 | 6 |
| How can more sustainable reusable equipment safely be used during and around the time of an operation? | 16 | 5.5 | 7 |
| How do we measure the carbon footprint of an operation? | 14 | 10 | 8 |
| How can healthcare professionals who deliver care during and around the time of an operation be encouraged to adopt sustainable actions in practice? | 12 | 13 | 9.5 |
| How can energy usage within an operating theatre be safely reduced? | 8 | 17 | 9.5 |
| How can we compare the environmental impacts of reusable and single-use equipment used during and around the time of an operation? | 10 | 17 | 11 |
| How can healthcare organisations more sustainably procure (obtain) medicines, equipment and items used during and around the time of an operation? | 9 | 21 | 12.5 |
| Can alternative, more environmentally sustainable, methods of disposal be used for waste that is generated during and around the time of an operation? | 6 | 24 | 12.5 |
| How can environmental sustainability be incorporated into the organisational management of operating theatres? | 15 | 17 | 14.5 |
| What are the best ways to educate healthcare professionals who provide care before, during and after operations, about sustainable healthcare? | 22 | 10 | 14.5 |
| What is the most sustainable way of providing equipment packs for an operation? | 19.5 | 13 | 16.5 |
| What is the most sustainable way to sterilise equipment used during an operation? | 19.5 | 13 | 16.5 |
| Can more efficient use of operating theatres and associated practices reduce the environmental impact of operations? | 27.5 | 10 | 18 |
| How can the waste of drugs be avoided during and around the time of an operation? | 21 | 17 | 19 |
| How can drug syringes be used better to reduce their environmental impact? | 12 | 26.5 | 20 |
| How can we increase the reuse and recycling of equipment used for rehabilitation in the recovery period after an operation? | 38.5 | 8 | 22 |
| What is the environmental impact of different anaesthetic techniques (e.g., different types of general, regional and local anaesthesia) used for the same operation? | 7 | 40.5 | 23.5 |
| How do we measure and compare the short- and long-term environmental impacts of surgical and non-surgical treatments for the same condition? | 35 | 17 | 26 |
| What are the barriers to using more sustainable anaesthetic practices? | 18 | 40.5 | 28.5 |
| How should the environmental impact of an operation be weighed against its clinical outcomes and financial costs? | 17 | 50.5 | 32 |

A total of 21 individuals attended the final prioritisation workshop, of whom eight classified themselves as patients, carers or members of the public, and 13 as healthcare professionals. The healthcare professionals comprised three surgeons, one operating department practitioner, five anaesthetists, one medical student, one foundation doctor, one optometrist, and one sustainability officer. Three of the patient, carer and public representatives were also members of the Greener Operations steering group. We noted that four of the patient, carer and public representatives had worked in healthcare at some point in their careers. Five observers from stakeholder organisations (e.g., the National Institute for Health and Care Research, Greener NHS) and the Greener Operations project leads and information specialists were present but not did not take part in the prioritisation discussions. The 25 indicative questions were ranked, and the top 10 priorities for research into sustainable peri-operative practice were agreed (table 3). All decisions were reached by consensus, with no majority votes required.

**Table 3:** Ranked research priorities from the final Greener Operations Priority setting workshop. The ‘Top 10’ are highlighted in green.

| Rank | Question |
| --- | --- |
| 1 | How can more sustainable reusable equipment safely be used during and around the time of an operation? |
| 2 | How can healthcare organisations more sustainably procure (obtain) medicines, equipment and items used during and around the time of an operation? |
| 3 | How can healthcare professionals who deliver care during and around the time of an operation be encouraged to adopt sustainable actions in practice? |
| 4 | Can more efficient use of operating theatres and associated practices reduce the environmental impact of operations? |
| 5 | How can the amount of waste generated during and around the time of an operation be minimised? |
| 6 | How do we measure and compare the short- and long-term environmental impacts of surgical and non-surgical treatments for the same condition? |
| 7 | What is the environmental impact of different anaesthetic techniques (e.g., different types of general, regional and local anaesthesia) used for the same operation? |
| 8 | How should the environmental impact of an operation be weighed against its clinical outcomes and financial costs? |
| 9 | How can environmental sustainability be incorporated into the organisational management of operating theatres? |
| 10 | What are the most sustainable forms of effective infection prevention and control used around the time of an operation (e.g., PPE, drapes, clean air ventilation)? |
| 11 | How do we measure the carbon footprint of an operation? |
| 12 | What are the best ways to educate healthcare professionals who provide care before, during and after operations, about sustainable healthcare? |
| 13 | Can equipment be recycled or repaired, instead of being disposed of, to reduce its environmental impact? |
| 14 | How can the waste of drugs be avoided during and around the time of an operation? |
| 15 | What alternative, more sustainable, materials can replace plastic packaging and equipment used during and around the time of an operation? |
| 16 | How do we define and avoid low-benefit or unnecessary operations? |
| 17 | How can energy usage within an operating theatre be safely reduced? |
| 18 | What are the barriers to using more sustainable anaesthetic practices? |
| 19 | How can we increase the reuse and recycling of equipment used for rehabilitation in the recovery period after an operation? |
| 20 | Can alternative, more environmentally sustainable, methods of disposal be used for waste that is generated during and around the time of an operation? |
| 21 | What is the most sustainable way of providing equipment packs for an operation? |
| 22 | How can we compare the environmental impacts of reusable and single-use equipment used during and around the time of an operation? |
| 23 | How much recyclable waste generated during an operation is being appropriately recycled? |
| 24 | What is the most sustainable way to sterilise equipment used during an operation? |
| 25 | How can drug syringes be used better to reduce their environmental impact? |

## Discussion

The Greener Operations PSP has identified the top 25 research priorities for sustainable peri-operative practice, with an emphasis on the top 10. This provides a robust basis for end-user focussed research into mitigating the environmental impacts of a resource-intensive area of healthcare at a time of climate crisis.^4^ Despite a recent increase in the number of publications into sustainable healthcare in the peri-operative period,^22^ this remains a relatively under-investigated area - as indicated by our literature review that revealed no ‘completely answered’ indicative questions. Though there are established sustainability measures that are already being implemented at scale, research will be required to understand how a fully sustainable healthcare system can be achieved.^4^ Furthermore, implementation research will be required to identify how to achieve some of the behavioural elements (e.g., changes in practice) that have been identified as important but not yet integrated into practice. The top 10 research priorities relate to research uncertainties across multiple areas of research interest, including implementation (priorities 1 and 3), manufacturing and supply (priority 2), management (priorities 4 and 9), waste (priorities 1 and 5), surgery (priority 6), anaesthesia (priority 7), medical ethics (priority 8), economics (priority 8 and 9) and infection control (priority 10). This both underlines the interdisciplinary relevance of the PSP and highlights the complexity of the sustainability challenge faced by healthcare.^23^

Our PSP had an above average overall number of suggestions in the primary survey (1,926, compared to the mean of 1,723),^24^ with each respondent contributing more than six suggestions on average. This is likely to be representative of the enthusiasm for this area of study amongst the participants. Whilst the JLA process aims to engage a broad range of respondents, it is not uncommon for PSPs to have an imbalance in the background of survey respondents.^25-28^ Our PSP had a preponderance of healthcare professional respondents, with 78.2% fitting into this category across both surveys. Our use of internet-based approaches e.g., social media and online surveys, may have contributed to the imbalance of respondents, as the likelihood of having had an operation (therefore feeling more informed to comment on the peri-operative process) increases with age, whereas internet usage is inversely proportional to age.^29,30^ Methods to address this such as in-person or paper surveys (e.g., made available at patient encounters such as clinics as in other PSPs) were not feasible for our project given the restrictions on social contact owing to the COVID-19 pandemic at the time of the work. However, we are confident that the overall results were representative of both healthcare and non-healthcare groups, because the JLA methodology controls for imbalances in survey response numbers, and also because the results of the interim survey showed strong alignment in priorities between both groups (Table 2). This was further strengthened through active patient, carer and public participation in the final workshop.

The Greener Operations priorities should provide a valuable resource for researchers and funders. Based on our literature review, we are confident that none of the identified research priorities have been completely answered by existing research. However, some priorities have attracted a significant research effort in recent years and have therefore been partially answered. In particular, the ‘carbon footprints’ of various anaesthetic agents, disposable and re-usable instruments, infection control supplies, and PPE (relating to priorities 1, 7 and 10) have been investigated.^31-35^ Here, there is an increasing amount of coherent data on climate impacts (i.e., ‘carbon footprints’), but other aspects (e.g., the ecotoxic effects of plastic waste and / or drug and metabolite disposal) remain under-investigated.^35^ Furthermore, sustainable peri-operative care is an area of current innovation, and new developments may render current concepts rapidly outdated.^36,37^ Nevertheless, experts in the field who are aware of the current literature may consider some questions outside the Greener Operations rankings to be of greater priority than some of those within. The PSP process should not be seen to diminish the value of this expertise. Rather, it adds insight into what the end users of research – patients, carers, public and clinicians – perceive to be important about environmentally sustainable peri-operative care. The priority questions are intentionally broad in scope, and we encourage researchers to draw on them in the development of projects. Of note, there appears to be an increasing number of funding calls relevant to this topic area, which we hope will be maintained in the future.^36,38^

Greener Operations has identified the top 10 research priorities for sustainable peri-operative care as agreed by a wide range of healthcare professionals, patients, carers and members of the public. Our project has explored a priority area for healthcare and identified a diverse range of research topics for exploration and innovation that will benefit both the NHS and healthcare outside the UK.^4^ We hope that our work will be of use to researchers and funders, as part of an urgent and universal effort to achieve high-quality healthcare with minimal environmental harm. Greener Operations is the first PSP undertaken by the JLA in sustainable healthcare and, to our knowledge, the first research priority setting exercise carried out in any field of sustainable healthcare. In addition to agreeing priority research areas for investigation, we have demonstrated that a PSP focussed on sustainable healthcare is feasible. Given the pressing nature of the climate crisis, we hope that colleagues in other fields will draw on our experience to conduct further sustainability-related PSPs.

## Data Availability

All data produced in the present study are available upon reasonable request to the authors

## Acknowledgments

The authors wish to acknowledge the contributions of Suzannah Kinsella, Toto Gronlund, Patricia Ellis and Amy Street for facilitating the final priority setting workshop; Manchester University NHS Foundation Trust Library Services for supporting the literature reviews; the Manchester University NHS Foundation Trust Sustainability Team for supporting the administration of the PSP; and the Association of Anaesthetists for providing the venue for the final priority setting workshop.

The authors thank the partner organisations for supporting the PSP: Association of Anaesthetists, Association of Coloproctology of Great Britain and Ireland, Bay Wide Maternity Voices, British Anaesthetic and Recovery Nurses Association, British Association of Day Surgery, Centre for Perioperative Care, Centre for Sustainable Healthcare, College of Operating Department Practitioners, Faculty of Intensive Care Medicine, Green Health Wales, Greener Anaesthesia and Sustainability Project, Health Innovation Manchester, Intensive Care Society, Perioperative Care Collaboration, Prevent Breast Cancer, Regional Anaesthesia UK, Royal College of Anaesthetists, Royal College of Nursing Perioperative Forum, Royal College of Obstetricians and Gynaecologists, Royal College of Ophthalmologists, Royal College of Surgeons of Edinburgh, Royal College of Surgeons of England, Society for Cardiothoracic Surgery in Great Britain and Ireland, Steps, Students Organising for Sustainability, UK Health Alliance on Climate Change, UK Clinical Pharmacy Association, and Ysbyty Gwynedd Green Group.

The authors thank the volunteers for participating in the final workshop: Yesmin Begum, Mood Bhutta, Claire Cruikshanks, James Dalton, Jenny Dorey, Bob Evans, Janice Fazackerley, Elizabeth Fitzhugh, Brooke Gerrie, Jean Hinton, John Hitchman, Rachel Kearns, Pinky Kotecha, Adam Peckham-Cooper, Carol Pellowe, Rebecca Pugsley, Meghan Richold, Patricia Romero, Kaushik Shah, Jennifer Tempany, and Katie Whitehouse.

## Author’s contributions

All authors made substantial contributions to the conception and design of the work; HN, MCS and OS contributed to data acquisition; all authors contributed to the analysis and interpretation of data. HN, MCS and CS drafted the work; all authors revised it critically for important intellectual content. All authors have approved the final version for publication and agree to be accountable for all aspects of the work in ensuring that questions related to the accuracy or integrity of any part of the work are appropriately investigated and resolved. David Riding made substantial contributions to the conception and design of the work and the analysis and interpretation of data as a member of the Steering Group but does not meet the ICMJE criteria for authorship; the authors thank him for his valuable contributions to the work.

## References

1. Sherman JD, McGain F, Lem M, Mortimer F, Jonas WB, MacNeill AJ. Net zero healthcare: a call for clinician action. BMJ. 2021;374:1323.

2. Watts N, Amann M, Arnell N, et al. The 2020 report of the Lancet Countdown on health and climate change: responding to converging crises. Lancet. 2021;397:129–70.

3. Reed S, Göpfert A, Wood S, Allwood D, Warburton W. Building healthier communities: the role of the NHS as an anchor institution. London: Health Foundation 2019.

4. Greener NHS. Delivering a ‘Net Zero’ National Health Service. London: NHS England and NHS Improvement 2020.

5. White SM, Shelton CL, Gelb AW, et al. Principles of environmentally-sustainable anaesthesia: a global consensus statement from the World Federation of Societies of Anaesthesiologists. Anaesthesia. 2022;71:201–12.

6. Chakera A, Fennell-Wells A, Allen C. Piped nitrous oxide waste reduction strategy. 2021 https://anaesthetists.org/Portals/0/PDFs/Environment/Nitrous%20waste%20methodology.pdf (accessed: 6th June 2022).

7. General Medical Council. Tomorrow’s Doctors. London: General Medical Council 2018.

8. Nursing and Midwifery Council. Standards of Proficiency for Midwives. London: Nursing and Midwifery Council 2019.

9. Royal College of Anaesthetists. 2021 Curriculum Learning Syllabus: Stage 1. 2021. https://rcoa.ac.uk/documents/2021-curriculum-learning-syllabus-stage-1/introduction (accessed: 6th June 2022).

10. Holmer H, Bekele A, Hagander L, et al. Evaluating the collection, comparability and findings of six global surgery indicators. Br J Surg. 2019;106:e138–e150.

11. Meara JG, Leather AJ, Hagander L, et al Global Surgery 2030: evidence and solutions for achieving health, welfare, and economic development. Lancet. 2015;386:569–624.

12. MacNeill AJ, Lillywhite R, Brown CJ. The impact of surgery on global climate: a carbon footprinting study of operating theatres in three health systems. Lancet Planet Health. 2017;1:e381–e388.

13. Hutchins DC, White SM. Coming round to recycling. BMJ. 2009;338:b609.

14. Rizan C, Steinbach I, Nicholson R, et al. The carbon footprint of surgical operations: a systematic review. Ann Surg. 2020;272:986–995.

15. Lawson C. Fellowship in Environmentally Sustainable Anaesthesia. Anaesthesia News 2019;379:5–6.

16. Centre for Sustainable Healthcare. Green Surgery Challenge. 2022. https://sustainablehealthcare.org.uk/what-we-do/green-surgery-challenge (accessed 9th June 2022)

17. Publications – something that docuements increase – maybe just a pubmed search record?

18. James Lind Alliance. About the James Lind Alliance. 2022. https://www.jla.nihr.ac.uk/about-the-james-lind-alliance/ (accessed 28th June 2022)

19. James Lind Alliance. The PSPs. 2022. https://www.jla.nihr.ac.uk/priority-setting-partnerships/ (accessed 28th June 2022)

20. James Lind Alliance. The James Lind Alliance Guidebook, Version 10. Southampton: James Lind Alliance 2021

21. Greener Operations Steering Group. Greener Operations: Sustainable Peri-Operative Practice PSP Protocol. 2021. https://www.jla.nihr.ac.uk/documents/greener-operations-sustainable-peri-operative-practice-psp-protocol/27106 (accessed 26th June 2022)

22. PubMed. (sustain*) AND (enviro*) AND ((peri-operative) or perioperative) - Search Results. 2022. https://pubmed.ncbi.nlm.nih.gov/?term=%28sustain*%29+AND+%28enviro*%29+AND+%28%28peri-operative%29+or+perioperative%29 (accessed 28th June 2022).

23. Stancliffe R. 10 years to green the NHS and the health sector. Lancet Planet Health. 2020;4:e126–e127

24. Nygaard A, Halvorsrud L, Linnerud S, et al The James Lind Alliance process approach: scoping review. BMJ Open 2019;9:e027473

25. Bretherton CP, Claireaux HA, Gower J, et al Research priorities for the management of complex fractures: a UK priority setting partnership with the James Lind Alliance BMJ Open 2021;11:e057198

26. Karantana A, Davis T, Kennedy D, et al Common hand and wrist conditions: creation of UK research priorities defined by a James Lind Alliance Priority Setting Partnership BMJ Open 2021;11:e044207

27. Lai FY, Abbasciano RG, Tabberer B Steering Group of the James Lind Alliance Heart Surgery Priority Setting Partnership, et al Identifying research priorities in cardiac surgery: a report from the James Lind Alliance Priority Setting Partnership in adult heart surgery BMJ Open 2020;10:e038001

28. Morris C, Simkiss D, Busk M, et al Setting research priorities to improve the health of children and young people with neurodisability: a British Academy of Childhood Disability-James Lind Alliance Research Priority Setting Partnership BMJ Open 2015;5:e006233

29. Fowler AJ, Abbott TEF, Prowle J, et al. Age of patients undergoing surgery. Br J Surg 2019;106:1012–8.

30. Office of National Statistics. Exploring the UK’s digital divide. 2019, https://www.ons.gov.uk/peoplepopulationandcommunity/householdcharacteristics/homeinternetandsocialmediausage/articles/exploringtheuksdigitaldivide/2019-03-04 (accessed 29th June 2022)

31. Eckelman M, Mosher M, Gonzalez A, Sherman J. Comparative life cycle assessment of disposable and reusable laryngeal mask airways. Anesth Analg. 2012;114:1067–72

32. Rizan C, Bhutta MF. Environmental impact and life cycle financial cost of hybrid (reusable/single-use) instruments versus single-use equivalents in laparoscopic cholecystectomy. Surg Endosc. 2022 Jun;36:4067–4078

33. Rizan C, Reed M, Bhutta MF. Environmental impact of personal protective equipment distributed for use by health and social care services in England in the first six months of the COVID-19 pandemic. J R Soc Med. 2021;114:250–263

34. Narayanan H, Raistrick C, Pierce JMT, Shelton C. Carbon footprint of inhalational and total intravenous anaesthesia for paediatric anaesthesia: a modelling study. Br J Anaesth. 2022;doi: 10.1016/j.bja.2022.04.022

35. McGain F, Muret J, Lawson C, Sherman JD. Environmental sustainability in anaesthesia and critical care. Br J Anaesth. 2020;125:680–692

36. SBRI Healthcare. Delivering a Net Zero NHS: Competition for Development Funding. London: NHS England, NHS Improvement and SBRI Healthcare 2021.

37. Devlin-Hegedus JA, McGain F, Harris RD, Sherman JD. Action guidance for addressing pollution from inhalational anaesthetics. Anaesthesia. 2022;doi:10.1111/anae.15785

38. National Institute for Health and Care Research. Delivering a Sustainable Health and Care System - specification document. 2022. https://www.nihr.ac.uk/documents/delivering-a-sustainable-health-and-care-system/29647 (accessed 29th June 2022)

